# Development and Validation of a Survival Calculator for Hospitalized Patients with COVID-19

**DOI:** 10.1101/2020.04.22.20075416

**Authors:** Todd J. Levy, Safiya Richardson, Kevin Coppa, Douglas P. Barnaby, Thomas McGinn, Lance B. Becker, Karina W. Davidson, Stuart L. Cohen, Jamie S. Hirsch, Theodoros P. Zanos, And the Northwell & Maimonides COVID-19 Research Consortium, Hannah Bodenstein, Shubham Debnath, Andrew J. Dominello, Louise Falzon, Michael Gitman, Jay M. Goldstein, Crystal Herron, Eun-Ji Kim, Lawrence Lau, Zachary S. Lockerman, Alexander Makhnevich, Jazmin N. Mogavero, Ernesto P. Molmenti, Marc d. Paradis, Viktor Tóth

## Abstract

**Background:** Chinese studies reported predictors of severe disease and mortality associated with coronavirus disease 2019 (COVID-19). A generalizable and simple survival calculator based on data from US patients hospitalized with COVID-19 has not yet been introduced.

**Objective:** Develop and validate a clinical tool to predict 7-day survival in patients hospitalized with COVID-19.

**Design:** Retrospective and prospective cohort study.

**Setting:** Thirteen acute care hospitals in the New York City area.

**Participants:** Adult patients hospitalized with a confirmed diagnosis of COVID-19. The development and internal validation cohort included patients hospitalized between March 1 and May 6, 2020. The external validation cohort included patients hospitalized between March 1 and May 5, 2020.

**Measurements:** Demographic, laboratory, clinical, and outcome data were extracted from the electronic health record. Optimal predictors and performance were identified using least absolute shrinkage and selection operator (LASSO) regression with receiver operating characteristic curves and measurements of area under the curve (AUC).

**Results:** The development and internal validation cohort included 11□095 patients with a median age of 65 years [interquartile range (IQR) 54-77]. Overall 7-day survival was 89%. Serum blood urea nitrogen, age, absolute neutrophil count, red cell distribution width, oxygen saturation, and serum sodium were identified as the 6 optimal of 42 possible predictors of survival. These factors constitute the NOCOS (Northwell COVID-19 Survival) Calculator. Performance in the internal validation, prospective validation, and external validation were marked by AUCs of 0.86, 0.82, and 0.82, respectively.

**Limitations:** All participants were hospitalized within the New York City area.

**Conclusions:** The NOCOS Calculator uses 6 factors routinely available at hospital admission to predict 7-day survival for patients hospitalized with COVID-19. The calculator is publicly available at https://feinstein.northwell.edu/NOCOS.

**Trial registration:** N/A

**Funding Source:** This work was supported by grants R24AG064191 from the National Institute on Aging, R01LM012836 from the National Library of Medicine, and K23HL145114 from the National Heart Lung and Blood Institute.

## INTRODUCTION

The World Health Organization designated coronavirus disease 2019 (COVID-19) as a global pandemic on March 11, 2020, with over 1 million confirmed cases worldwide (1). Estimates of severe disease range from 20% to 30%, and fatality rates range from 2% to 7% (2, 3). As healthcare facilities around the world struggle to provide care for rising numbers of critically ill patients, evidence-based tools to assist with prognosis and estimating disease severity are becoming increasingly important (4). These tools can guide conversations with patients and families, advise therapeutic decisions (e.g., admission to the intensive care unit), and align treatment plans with the likelihood of benefit (5).

Some clinical prediction tools have been established to estimate survival in patients with pneumonia or hospitalized with severe illness, including the Sequential Organ Failure Assessment (SOFA) and CURB-65 Scores. However, these tools have not been validated in patients with COVID-19. A recent study reported a clinical risk score for hospitalized patients with this disease (6). While the score relies on data routinely available at admission, it uses unstructured data, cannot predict survival alone, and is based on a cohort of patients hospitalized in China. Several other models predict outcomes in patients with COVID-19. However, many of these models are not peer-reviewed and are at a high risk of bias because of non-representative samples of control patients (7).

Our objective was to develop and externally validate a clinical prediction tool to estimate 7-day survival in patients hospitalized with COVID-19 in the United States. For this tool, we aimed to use exclusively discrete data points from the electronic health record (EHR), forgoing symptom-related records and radiology reads. By including only objective, structured data points that are routinely available at hospital admission, we could reduce ambiguity, improve external performance, and ensure that the tool could be used in most acute-care settings.

## METHODS

### Study Design

This study includes a retrospective analysis for development, retrospective internal validation, prospective internal validation, and external validation of a model to predict survival of patients hospitalized with COVID-19. The development cohort included patients admitted to 11 of 12 acute care facilities in the Northwell Health system between March 1 and April 23, 2020. The internal retrospective validation cohort included patients admitted to the remaining acute care tertiary facility in the Northwell Health System, Long Island Jewish Medical Center, between March 1 and May 7, 2020. Long Island Jewish Medical Center has the largest number of hospitalized patients with COVID-19 in the Northwell Health system. The internal prospective validation cohort included patients admitted to all 12 acute care facilities in the Northwell Health system between April 24 and May 6, 2020. The external validation cohort included patients admitted to Maimonides Medical Center, an affiliate of the Northwell Health system, between March 1 and May 12, 2020. The final date of follow-up was May 7, 2020 for the internal validation cohorts and May 12, 2020 for the external validation cohort (Figure 1A).

**Figure 1.**
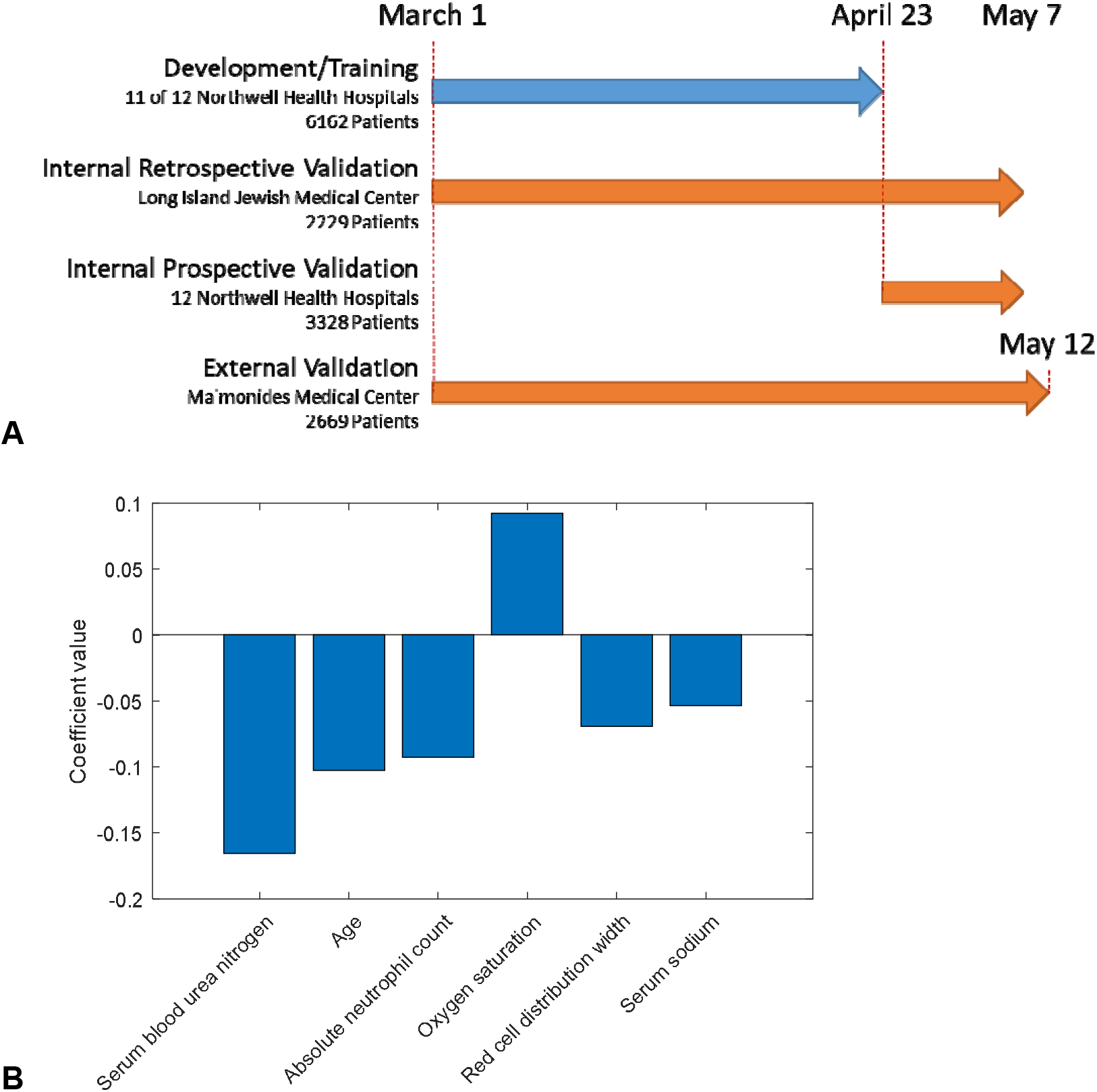
Study design and predictive performance of the NOCOS Calculator. (A) Training and validation datasets used to create and test the predictive performance of the NOCOS Calculator. The datasets include internal retrospective, internal prospective, and external datasets comprising 13□764 patients. Note that the internal retrospective and prospective validation sets overlap by 624 patients admitted to Long Island Jewish Medical Center. (B) Importance of the 6 predictors captured by the coefficients of the NOCOS Calculator. NOCOS = Northwell COVID-19 Survival.

Patients were included if they were adults (≥ 18 years old) admitted to the hospital with COVID-19 confirmed by a positive result from polymerase chain reaction testing of a nasopharyngeal sample. Clinical outcomes (i.e., discharges, mortality, length of stay) were monitored until the final date of follow-up. Patients were excluded if they received invasive mechanical ventilation before inpatient admission, either before presentation to or during their stay in the emergency department. Patients were also excluded if their length of stay was less than 7 days and they were still hospitalized on the final date of follow-up.

This study was approved by the Institutional Review Boards at Northwell Health and Maimonides Medical Center as minimal-risk research that used data collected for routine clinical practice, and as such, waived the requirement for informed consent.

### Data Acquisition

Data were collected from the enterprise electronic health record (EHR; Sunrise Clinical Manager, Allscripts, Chicago, IL). Transfers from 1 in-system hospital to another were merged and considered 1 hospital visit. Data collected for the development and internal validation of the tool included patient demographic information, comorbidities, laboratory values, and outcomes (i.e., death, length of stay, discharge). Data collected for the external validation included only the 6 predictor variables found in the development and internal validation process, length of stay, and final outcome (i.e., death, discharge).

### Potential Predictive Variables

Potential predictive variables were included if they were available for more than half of study patients at the time of admission. This approach ensured that the results would contain data points routinely available at admission. Continuous variables are presented as median and interquartile range (IQR), and categorical variables are expressed as the number of patients and percentage.

Demographic variables included age, gender, race, ethnicity, and language preference as English or non-English. Vitals signs included systolic blood pressure, diastolic blood pressure, heart rate, respiratory rate, oxygen saturation, temperature, body mass index, height, and weight. Comorbidities included coronary artery disease, diabetes, hypertension, heart failure, lung disease, and kidney disease. Laboratory variables included white blood cell count, absolute neutrophil count, automated lymphocyte count, automated eosinophil count, automated monocyte count, hemoglobin, red cell distribution width, automated platelet count, serum sodium, serum potassium, serum chloride, serum carbon dioxide, serum blood urea nitrogen, serum creatinine, estimated glomerular filtration rate, serum glucose, serum albumin, serum bilirubin, serum alkaline phosphatase, alanine aminotransferase, aspartate aminotransferase, and lactate.

### Outcomes

Outcomes collected included death, length of stay, and discharge. The primary outcome was 7-day survival. Patients who were discharged alive on any hospital day before or on hospital day 7 were considered to have survived. Patients who were alive and still in-hospital on hospital day 7 were considered to have survived. Patients who died before or on hospital day 7 were considered to have expired. Patients who were alive and still in-hospital at the study endpoint with a length of stay less than 7 days were excluded from the study.

### Prediction Model Development

The model was developed by analyzing 42 potential predictors for the patients hospitalized in 11 of 12 hospitals within the Northwell Health system and discharged on or before April 23, 2020 (Figure 1A). Patients hospitalized at Long Island Jewish Medical Center or discharged after April 23 were used for the internal retrospective and internal prospective validations, respectively. Least Absolute Shrinkage and Selection Operator (LASSO) regression was used to identify predictors that, when linearly combined, predict the survival of hospitalized patients with COVID-19 (8). Missing measurements were imputed using mean imputation. All analyses were performed in MATLAB 2019b (The Mathworks, Inc., Natick, MA).

By including an L_1_-norm regularization term that promotes sparsity, LASSO regression determines a subset of measurements in which only the strongest predictors remain in the model. The magnitudes of the coefficients relate to the predictive values of the normalized measurements, while coefficients of non-predictive measurements converge exactly to 0. The data are normalized by taking the z-score, which puts all measurements on the same scale. The mean and standard deviation of the measurements with coefficients that are not 0 are stored during training and applied to test data.

The training set was evaluated with the model using 50-fold cross-validation to prevent overfitting. The class-conditional likelihood functions of the LASSO predictions for survival past 7 days and expiration before 7 days were estimated, and the posterior probability of survival past 7 days was evaluated using Bayes Theorem. The regularization factor λ is a hyperparameter that was swept over a range while evaluating the area under the receiver operating characteristic (ROC) curve. After optimizing for λ, the number of predictors was fixed at 6 inputs. The variables identified were used to construct the Northwell COVID-19 Survival (NOCOS) Calculator, available publicly at https://feinstein.northwell.edu/NOCOS.

### Prediction Model Validation

The generalizability of the NOCOS Calculator was validated with the retrospective cohort from Long Island Jewish Medical Center, the internal prospective cohort, and the external cohort. The predictive performance of the model was assessed at the time of admission and every 2 days within the hospitalization via ROC and precision recall (PR) curves and the Area Under the Curve (AUC). We also tested the predictive value of the SOFA Score and CURB-65 Score for pneumonia severity, and we compared the AUCs for each score using the nonparametric DeLong method (9, 10).

To determine the performance of survival predictions for all calculators, operating points can be established by choosing thresholds on the probability scores. We chose 3 operating points for each calculator and provided the numbers of true positives, true negatives, false positives, and false negatives, as well as the positive predictive value (PPV) and negative predictive value (NPV) for each calculator.

### Calculation of SOFA and CURB-65 Scores

In 2 of the test datasets, data were assessed with the SOFA and CURB-65 Scores. The SOFA Score numerically quantifies the severity of failed organs based on PaO_2_/FiO_2_ (11), the Glasgow Coma Scale, mechanical ventilation (yes/no), platelets, bilirubin, mean arterial pressure or administration of vasoactive agents, and creatinine. Because PaO_2_ was not captured for most patients, we used a formula with SpO_2_ and FiO_2_ (Appendix Table 4). For patients missing a Glasgow Coma Scale score, the patient’s mental status was assessed using nursing documentation of mental status and level of consciousness. Missing data on all other variables were limited and treated similar to our imputations. No patients were missing data on ventilation status. The CURB-65 score is another mortality risk score based on confusion, blood urea nitrogen, respiratory rate, blood pressure, and age. For this score, a patient’s confusion level was assessed using nursing documentation.

## RESULTS

### Patient Characteristics

A total of 11 919 adult patients were hospitalized at the 12 Northwell Health acute care facilities between March 1 and May 7, 2020. Of these patients, 360 (3.02%) were excluded because they were still in the hospital at the study end point with a length of stay less than 7 days; 460 (3.86%) were excluded because they were transferred to a hospital outside of the health system and their outcomes were unknown; and 4 (0.03%) were excluded because they expired but were not marked as discharged in the EHR. The remaining 11□095 (93.09%) patients were included in the development and internal validation cohort. These patients had a median age of 65 years [IQR 54-77], and 42% were female. Overall 7-day survival was 89%. At the study end point, 10□207 (92%) patients were discharged alive or expired and 888 (8%) were still in the hospital. Baseline characteristics of included patients are presented in Table 1.

**Table 1.**
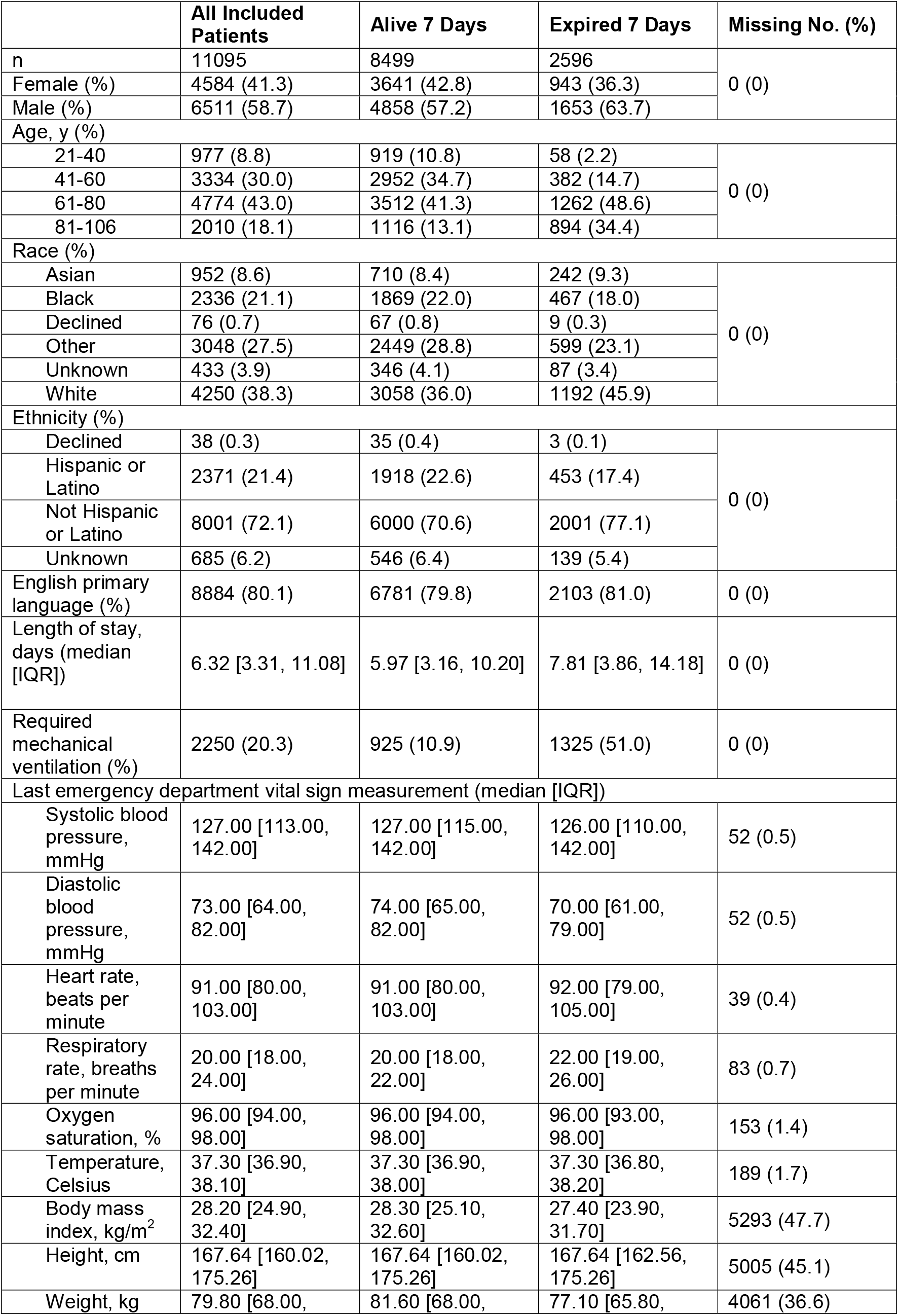
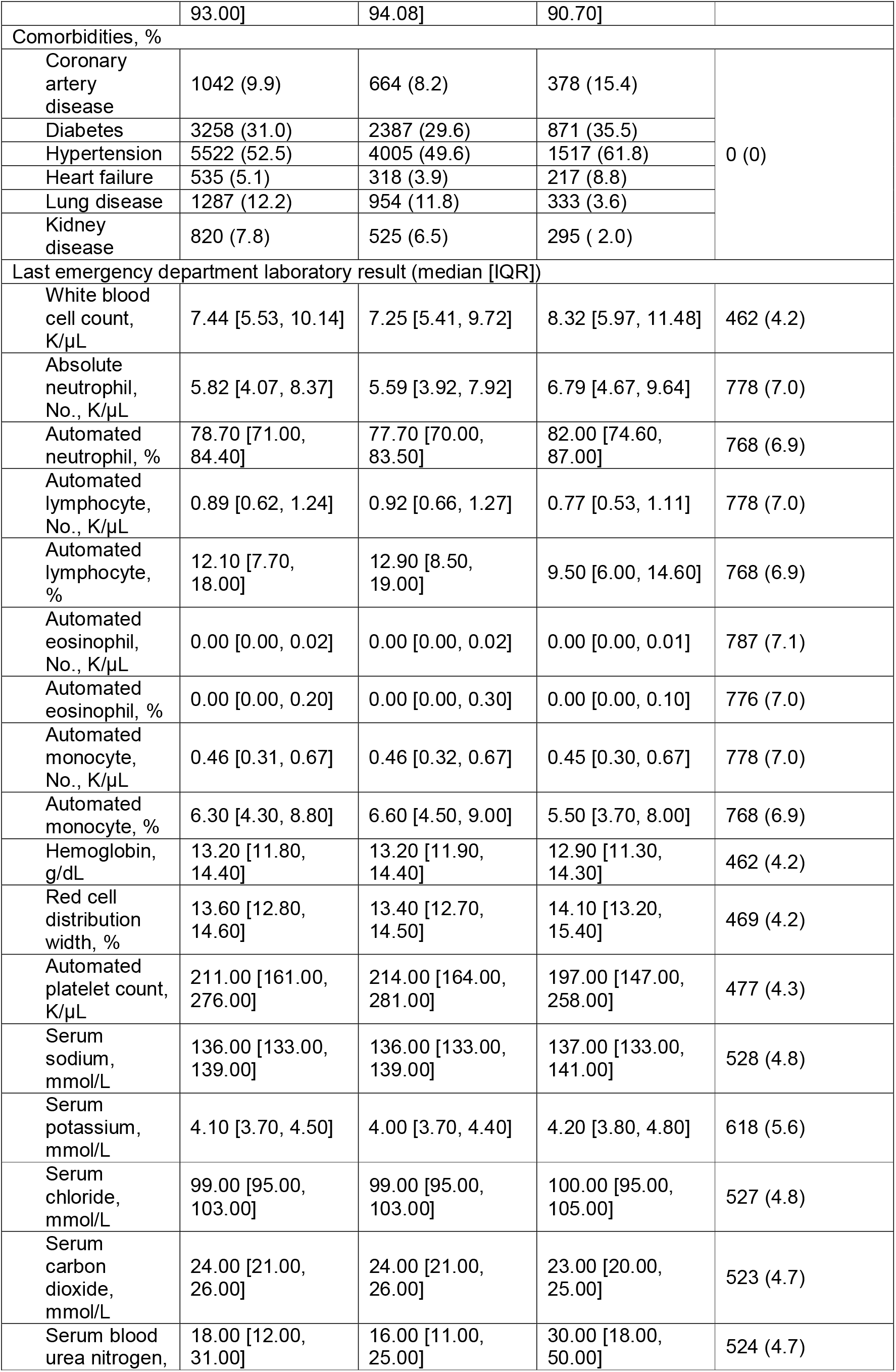
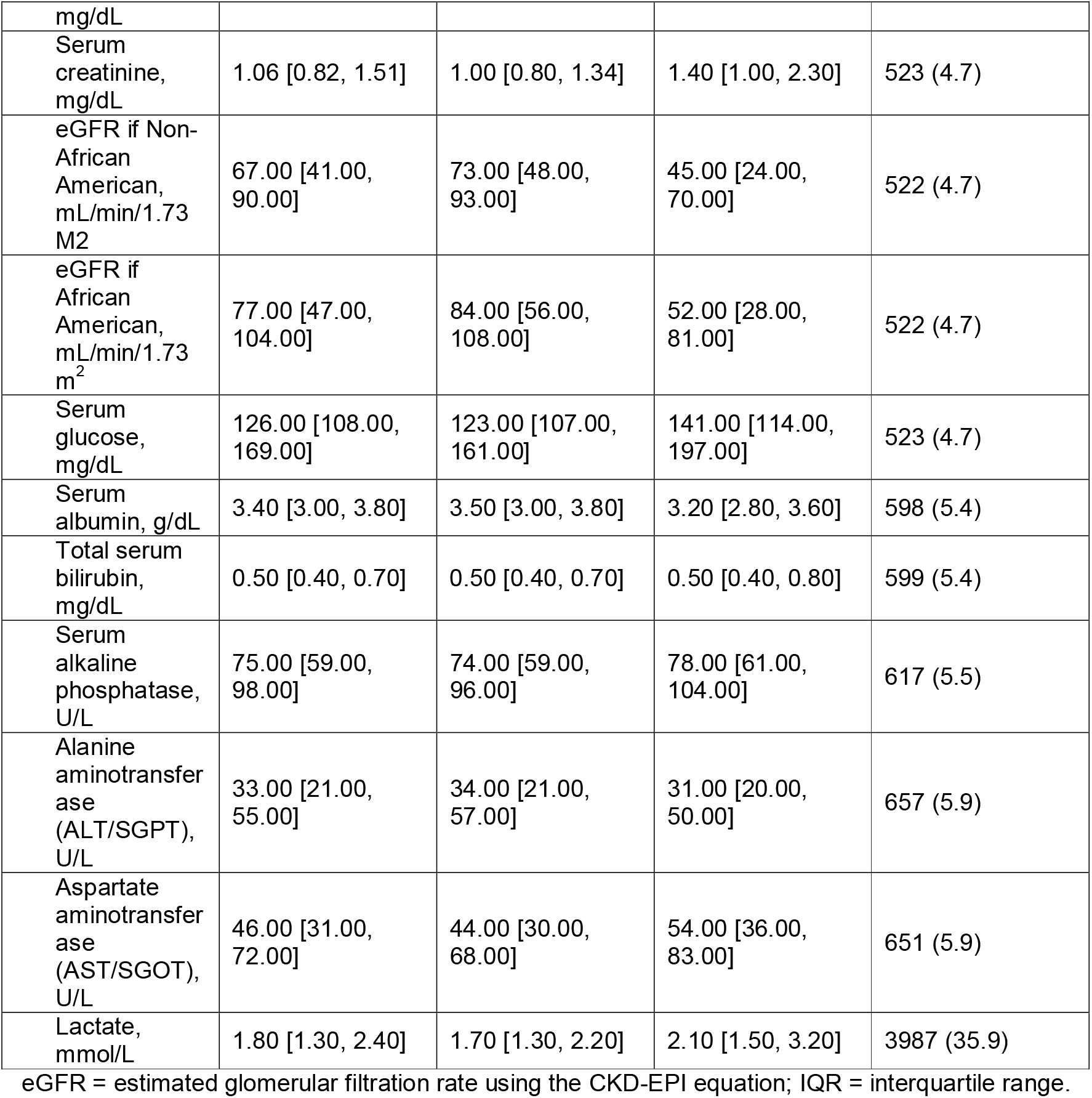
Demographic, Clinical, and Laboratory Data of All Patients Hospitalized at Northwell Health

### Survival Prediction Model

#### Development

To determine the predictors of survival, data were collected from patients hospitalized in 11 of 12 Northwell Health hospitals (*n* = 6162) (Figure 1A). The optimal predictors of survival, in decreasing order of relative predictive strength, were serum blood urea nitrogen, patient age, absolute neutrophil count, red cell distribution width, oxygen saturation, and serum sodium. For each predictor, the magnitude of the coefficient indicates the relative strength of the predictor in determining the outcome, and the sign of the coefficient corresponds to the sign of the correlation between the predictor and the outcome (Figure 1B).

#### Validation

For the internal retrospective validation, data were collected from patients hospitalized in Long Island Jewish Medical Center (*n* = 2229). These data were analyzed with NOCOS, and then the ROC and PR curves and AUC values were determined. The NOCOS Calculator resulted in an AUC of 0.86, which significantly outperformed the SOFA (AUC = 0.70; *p* < 0.05) and CURB-65 (AUC = 0.81; *p* < 0.05) Scores (Figure 2A).

**Figure 2.**
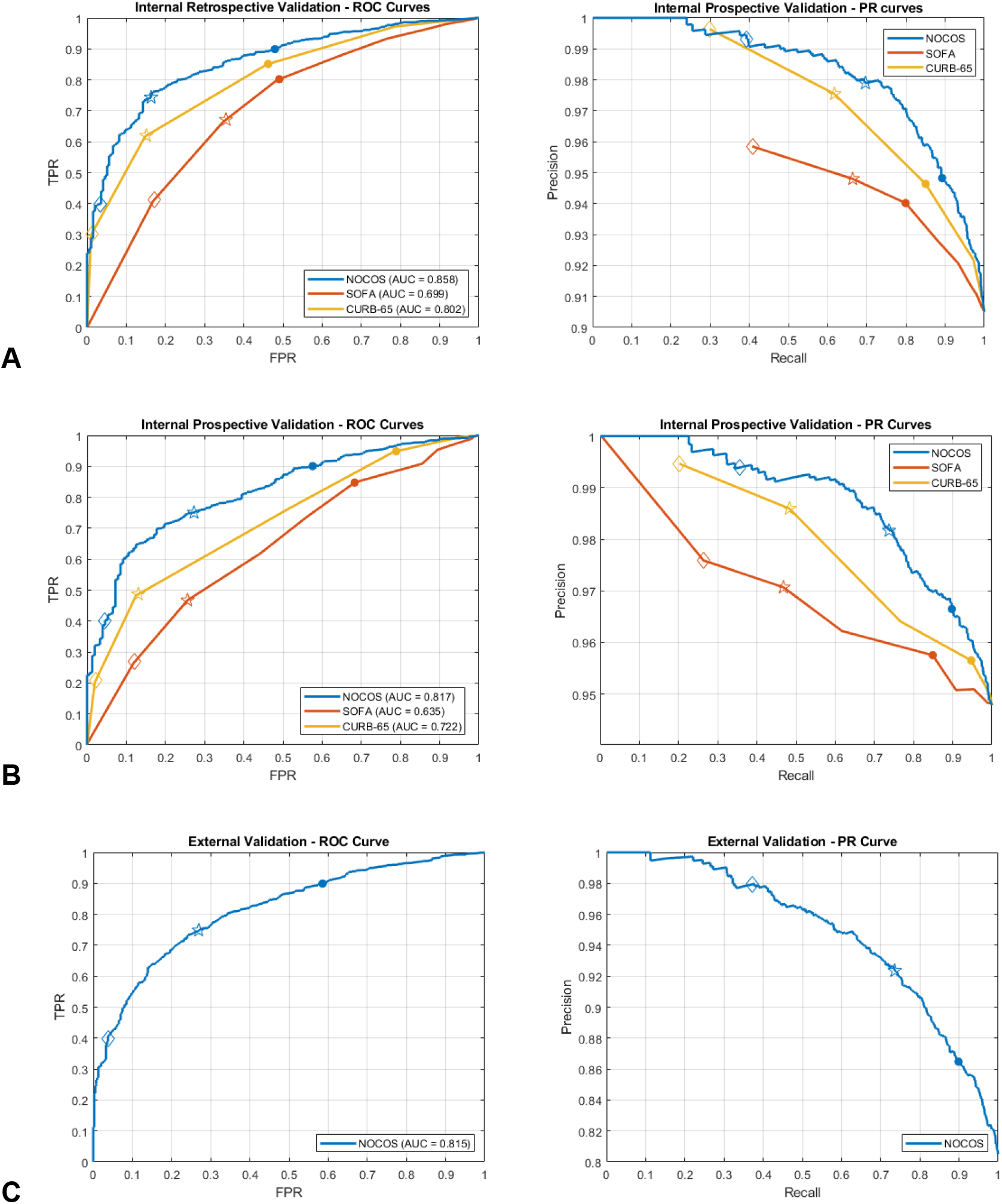
Predictive performance of the NOCOS Calculator on the internal retrospective, internal prospective, and external datasets. ROC and PR curves for the (A) internal retrospective validation with patients hospitalized at Long Island Jewish Hospital (*n* = 2229), (B) prospective validation with patients hospitalized across all 12 Northwell Health hospitals (*n* = 3328), and (C) external validation with patients hospitalized at Maimonides Medical Center (*n* = 2669). AUC = area under the curve; FPR = false positive rate; NOCOS = Northwell COVID-19 Survival; PR = precision recall; ROC = receiver operating characteristic; SOFA = Sequential Organ Failure Assessment; TPR = true positive rate.

For the internal prospective validation, data were collected from patients discharged from all 12 Northwell hospitals (*n* = 3328) (Figure 1A). Based on these data, the NOCOS Calculator (AUC = 0.82) significantly outperformed the SOFA (AUC = 0.64; *p* < 0.05) and CURB-65 (AUC = 0.72; *p* < 0.05) Scores (Figure 2B).

For the external validation, data were collected from patients hospitalized at Maimodines Medical Center (*n* = 2669) (Figure 1A). The NOCOS Calculator yielded a comparable AUC of 0.82 (Figure 2C); however, the SOFA and CURB-65 Scores were not readily available for this dataset because components of each score were not documented for all patients.

### Test Characteristics of the NOCOS Calculator, SOFA Score, and CURB-65 Score

To determine the performance of survival predictions for all calculators, operating points can be established by choosing thresholds on the probability scores. We chose 3 different operating points for each calculator for the internal retrospective validation (Long Island Jewish Medical Center) data and provided the number of true positives, true negatives, false positives, and false negatives. We also provided the PPV and NPV for each case (Table 2). In all cases, the NOCOS Calculator outperformed the SOFA and CURB-65 Scores. See Appendix Table 2 for metrics at the operating points of data from the internal retrospective validation (Long Island Jewish Medical Center), and see Appendix Table 3 for metrics from the external validation (Maimonides Medical Center).

**Table 2.** Confusion Matrices for Multiple Operating Points for the 3 Calculators Tested on Data from the Long Island Jewish Medical Center Dataset.

| **Operating Points A (●)** |  |  |  |
| --- | --- | --- | --- |
|  | **A1 (NOCOS)** | **A2 (SOFA)** | **A3 (CURB-65)** |
| Operating probability of survival/score | 51.6% | 3 | 2 |
| Predicted to survive & survived | 1667 | 1493 | 1588 |
| Predicted to expire & expired | 105 | 101 | 106 |
| Predicted to survive & expired | 91 | 95 | 90 |
| Predicted to expire & survived | 201 | 375 | 280 |
| Positive predictive value, % | 94.8 | 94.0 | 94.6 |
| Negative predictive value, % | 34.3 | 21.2 | 27.5 |
| **Operating Points B (★)** |  |  |  |
|  | **B1 (NOCOS)** | **B2 (SOFA)** | **B3 (CURB-65)** |
| Operating probability of survival/score | 75.4% | 2 | 1 |
| Predicted to survive & survived | 1301 | 1240 | 1153 |
| Predicted to expire & expired | 168 | 128 | 167 |
| Predicted to survive & expired | 28 | 68 | 29 |
| Predicted to expire & survived | 567 | 628 | 715 |
| Positive predictive value, % | 97.9 | 94.8 | 97.5 |
| Negative predictive value, % | 22.9 | 16.9 | 18.9 |
| **Operating Points C (◇)** |  |  |  |
|  | **C1 (NOCOS)** | **C2 (SOFA)** | **C3 (CURB-65)** |
| Operating probability of survival/score | 93.1% | 1 | 0 |
| Predicted to survive & survived | 734 | 762 | 558 |
| Predicted to expire & expired | 191 | 163 | 194 |
| Predicted to survive & expired | 5 | 33 | 2 |
| Predicted to expire & survived | 1134 | 1106 | 1310 |
| Positive predictive value, % | 99.3 | 95.8 | 99.6 |
| Negative predictive value, % | 14.4 | 12.8 | 12.9 |

NOCOS = Northwell COVID-19 Survival; SOFA = Sequential Organ Failure Assessment.

The NOCOS Calculator can also be reevaluated with updated labs and vitals as the patient’s condition progresses. The performance of the NOCOS calculator remained relatively stable over 10 days (Figure 3).

**Figure 3.**
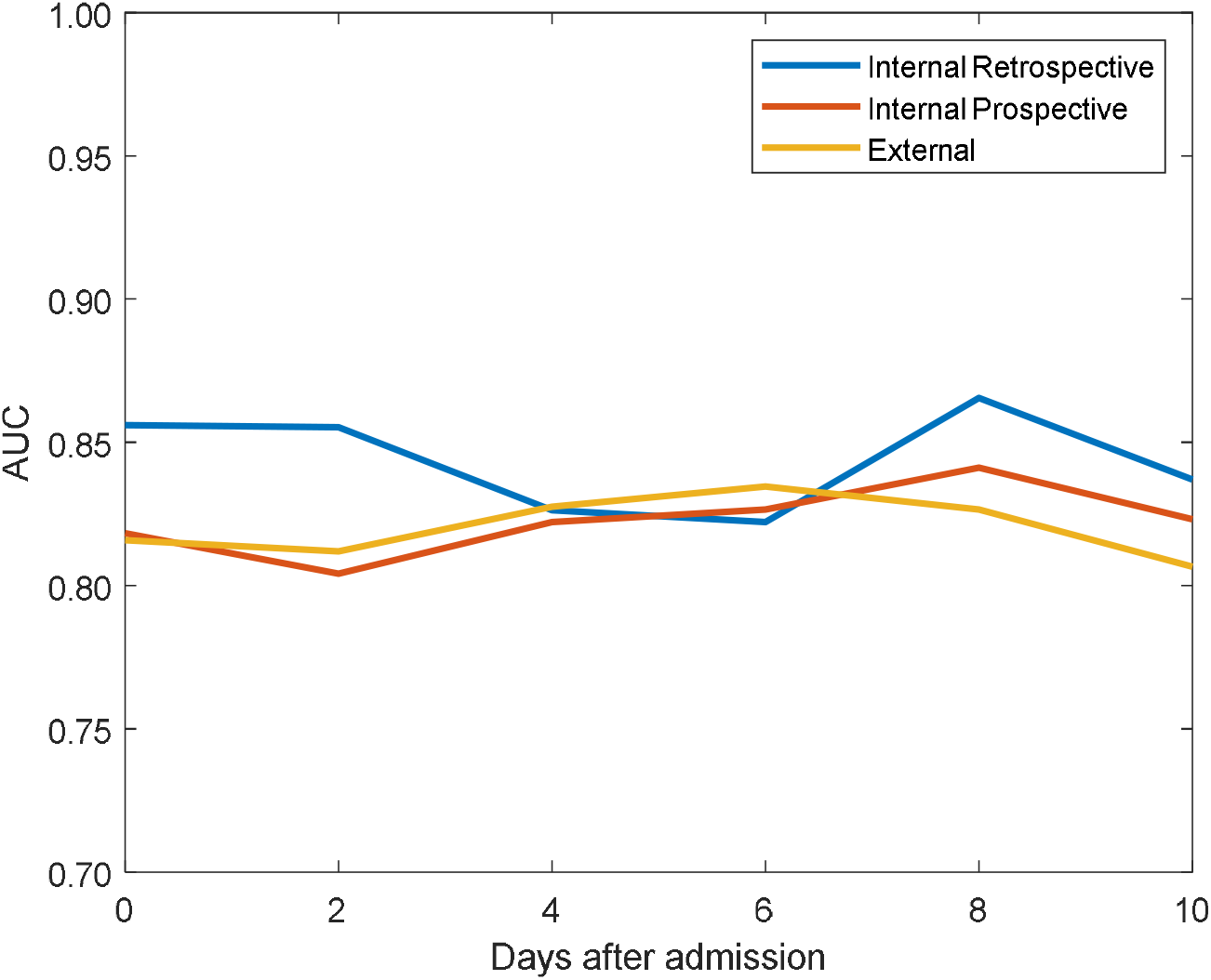
Stability of the predictive performance of the NOCOS Calculator across hospitalization days. AUC capturing the predictive performance of the NOCOS Calculator. The performance remained stable when the most recent values of the predictors were updated for up to 10 days during hospitalization after admission from the emergency department. AUC = Area Under the Curve; NOCOS = Northwell COVID-19 Survival.

## DISCUSSION

This study is the first to develop a model, the NOCOS Calculator, that predicts survival of patients hospitalized with COVID-19 in the United States. We created and validated the NOCOS Calculator with data on almost 14 000 patients, using only 6 clinical data points typically available to clinicians within the first 60 minutes of patient presentation. All data points are available as discrete inputs in most commercial EHRs, supporting that this calculator could be readily incorporated into tools to support clinical decisions. This calculator is publicly available at https://feinstein.northwell.edu/NOCOS and can be used by clinicians to estimate the probability of survival for their patients.

Several elements of the NOCOS Calculator have been either established as prognostic markers in other populations or identified as risk factors for severe illness or death in patients with COVID-19. Older age and elevated blood urea nitrogen (a marker of kidney dysfunction) have both been associated with increased mortality risk in patients with COVID-19.(12, 13) Hypoxemia, measured by lower levels of blood oxygen, has also been linked to increased mortality in this population (14). Neutrophil count, either individually or paired in a ratio with lymphocytes, also predicts disease severity in COVID-19 patients (15). While serum sodium has not yet been linked to COVID-19, it (16) has been independently and consistently associated with negative outcomes in other populations (17) and disease states.(18, 19) Elevated values of red cell distribution width often suggest chronic disease states and inflammation.(20, 21) An increased red cell distribution width may also be an effect of COVID-19 on iron displacement of the heme molecule, leading to impaired red blood cells, free-radical formation, and a toxic effect to the lungs.(22)

The NOCOS Calculator performs well with the 6 early measurements, and it retains its predictive performance as these measurements are updated over at least 10 days throughout the hospitalization of the patient (Figure 3). This finding supports that the most up-to-date values of the 6 measures can accurately predict survival. Moreover, while we present the calculator output as a probability score, a specific operating point can be chosen to provide a binary outcome prediction with significant accuracy. Stakeholders can choose an operating point, and local clinical teams can adjust thresholds toward a more stringent or risk-averse solution (Table 2) based on the rapidly changing needs during this pandemic.

Due to the challenges that arise during the ongoing COVID-19 pandemic, we need robust tools to aid in making complex clinical decisions. Using well-known clinical calculators, such as the SOFA or CURB-65 Scores, can be useful. However, these scores are limited by their accuracy and the ease of collecting necessary measurements to construct the scores. They also use input variables, such as confusion (for CURB-65) and the Glasgow Coma Scale (for SOFA), both of which are ambiguous, difficult to measure, and frequently unavailable, as shown by our external validation dataset. We found that the NOCOS Calculator consistently outperformed both the CURB-65 and SOFA scores in both our validation datasets.

### Limitations

The study population only included patients within the New York City metropolitan area. However, given the diverse demographic population of the region, we expect the model to generalize to patients at centers outside of this geographic area. The data were collected entirely from EHR reports, which supported robust and rapid analysis of a large cohort of patients. However, we did not include data elements that would require manual chart review. Due to the retrospective study design, not all laboratory tests were completed on all patients, and the performance of these variables could not be adequately assessed. To optimize for usability and portability, the analysis was designed to be linear and to include a minimum number of predictors. Non-linear or convolutional/recurrent models may provide improved performance but might not be easily used at all centers.

### Conclusion

This study is the first to develop and externally validate a simple predictive model of survival for hospitalized patients with COVID-19 based on structured, objective data that is routinely available at admission in the United States. Serum blood urea nitrogen, age, absolute neutrophil count, red cell distribution width, oxygen saturation, and serum sodium were identified as the 6 optimal predictors of survival. The NOCOS Calculator can predict survival more accurately than commonly used survival predictors.

## Data Availability

The data that support the findings of this study are available on request from. The data are not publicly available due to restrictions as it could compromise the privacy of research participants.

http://feinstein.northwell.edu/nocos

## Acknowledgments

We acknowledge and honor all our Northwell team members who consistently put themselves in harm’s way during the COVID-19 pandemic. We dedicate this article to them, as their vital contribution to knowledge about COVID-19 and sacrifices on the behalf of patients made it possible.

---

**Appendix Table 1.**
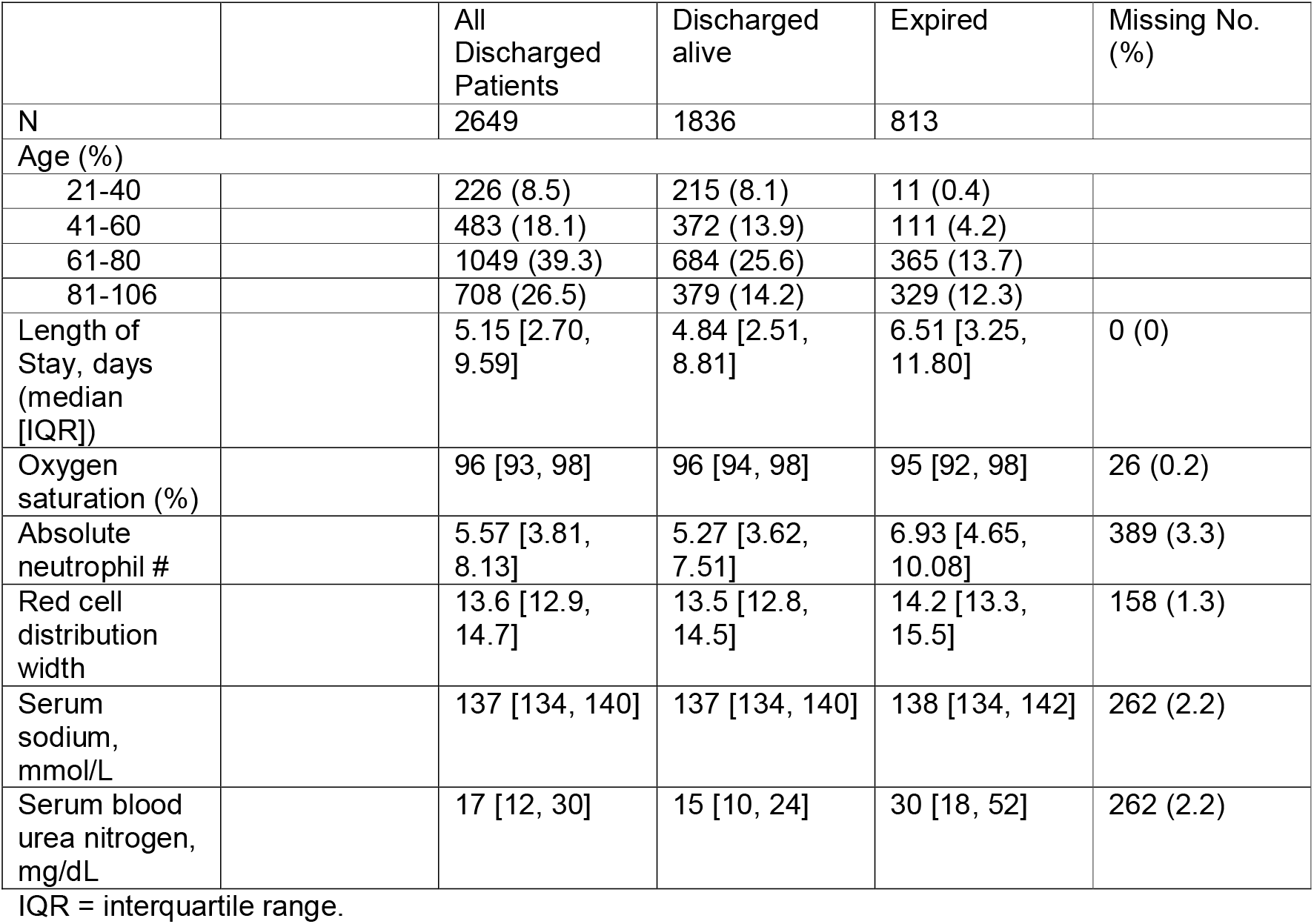
Summary of the 6 Predictor Variables from External Validation Data from Patients hospitalized at Maimonides Medical Center

|  |  | All Discharged Patients | Discharged alive | Expired | Missing No. (%) |
| --- | --- | --- | --- | --- | --- |
| N |  | 2649 | 1836 | 813 |  |
| Age (%) |  |  |  |  |  |
| 21-40 |  | 226 (8.5) | 215 (8.1) | 11 (0.4) |  |
| 41-60 |  | 483 (18.1) | 372 (13.9) | 111 (4.2) |  |
| 61-80 |  | 1049 (39.3) | 684 (25.6) | 365 (13.7) |  |
| 81-106 |  | 708 (26.5) | 379 (14.2) | 329 (12.3) |  |
| Length of Stay, days (median [IQR]) |  | 5.15 [2.70, 9.59] | 4.84 [2.51, 8.81] | 6.51 [3.25, 11.80] | 0 (0) |
| Oxygen saturation (%) |  | 96 [93, 98] | 96 [94, 98] | 95 [92, 98] | 26 (0.2) |
| Absolute neutrophil # |  | 5.57 [3.81, 8.13] | 5.27 [3.62, 7.51] | 6.93 [4.65, 10.08] | 389 (3.3) |
| Red cell distribution width |  | 13.6 [12.9, 14.7] | 13.5 [12.8, 14.5] | 14.2 [13.3, 15.5] | 158 (1.3) |
| Serum sodium, mmol/L |  | 137 [134, 140] | 137 [134, 140] | 138 [134, 142] | 262 (2.2) |
| Serum blood urea nitrogen, mg/dL |  | 17 [12, 30] | 15 [10, 24] | 30 [18, 52] | 262 (2.2) |

IQR = interquartile range.

**Appendix Table 2.**
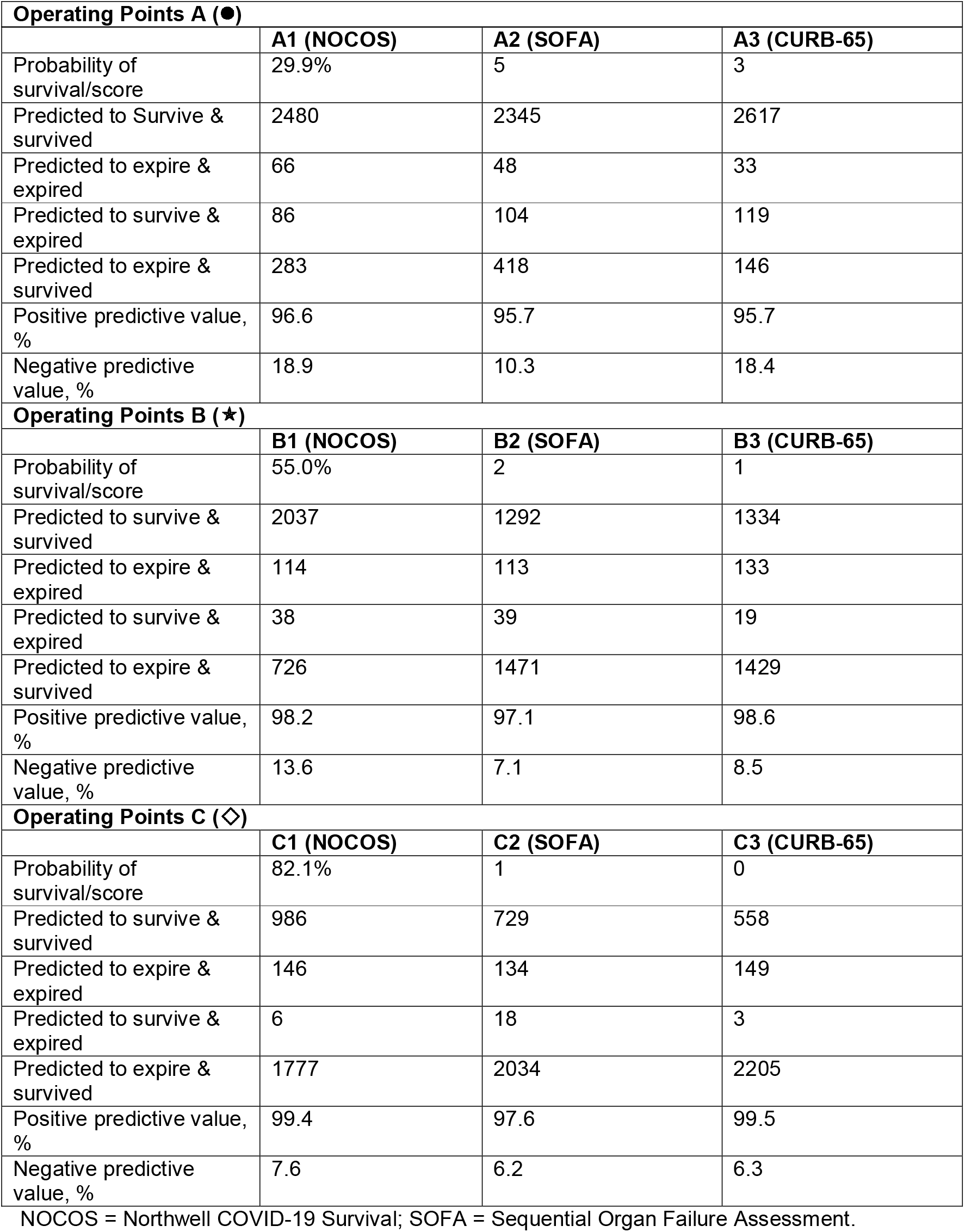
Confusion Matrices for Multiple Operating Points for the 3 Calculators Tested on the Prospective Dataset.

|  | **A1 (NOCOS)** | **A2 (SOFA)** | **A3 (CURB-65)** |
| --- | --- | --- | --- |
| Probability of survival/score | 29.9% | 5 | 3 |
| Predicted to Survive & survived | 2480 | 2345 | 2617 |
| Predicted to expire & expired | 66 | 48 | 33 |
| Predicted to survive & expired | 86 | 104 | 119 |
| Predicted to expire & survived | 283 | 418 | 146 |
| Positive predictive value, % | 96.6 | 95.7 | 95.7 |
| Negative predictive value, % | 18.9 | 10.3 | 18.4 |
| **Operating Points B (★)** |  |  |  |
|  | **B1 (NOCOS)** | **B2 (SOFA)** | **B3 (CURB-65)** |
| Probability of survival/score | 55.0% | 2 | 1 |
| Predicted to survive & survived | 2037 | 1292 | 1334 |
| Predicted to expire & expired | 114 | 113 | 133 |
| Predicted to survive & expired | 38 | 39 | 19 |
| Predicted to expire & survived | 726 | 1471 | 1429 |
| Positive predictive value, % | 98.2 | 97.1 | 98.6 |
| Negative predictive value, % | 13.6 | 7.1 | 8.5 |
| **Operating Points C (◇)** |  |  |  |
|  | **C1 (NOCOS)** | **C2 (SOFA)** | **C3 (CURB-65)** |
| Probability of survival/score | 82.1% | 1 | 0 |
| Predicted to survive & survived | 986 | 729 | 558 |
| Predicted to expire & expired | 146 | 134 | 149 |
| Predicted to survive & expired | 6 | 18 | 3 |
| Predicted to expire & survived | 1777 | 2034 | 2205 |
| Positive predictive value, % | 99.4 | 97.6 | 99.5 |
| Negative predictive value, % | 7.6 | 6.2 | 6.3 |

NOCOS = Northwell COVID-19 Survival; SOFA = Sequential Organ Failure Assessment.

**Appendix Table 3.** Confusion Matrices for Multiple Operating Points for the NOCOS Calculator Tested on the External Dataset.

| **Operating Points A (●)** |  |
| --- | --- |
|  | **A1 (NOCOS)** |
| Probability of survival | 36.6% |
| Predicted to survive & survived | 1490 |
| Predicted to expire & expired | 168 |
| Predicted to survive & expired | 233 |
| Predicted to expire & survived | 168 |
| Positive predictive value, % | 86.5 |
| Negative predictive value, % | 50.0 |
| **Operating Points B (★)** |  |
|  | **B1 (NOCOS)** |
| Probability of survival | 62.1% |
| Predicted to survive & survived | 1220 |
| Predicted to expire & expired | 300 |
| Predicted to survive & expired | 101 |
| Predicted to expire & survived | 438 |
| Positive predictive value, % | 92.4 |
| Negative predictive value, % | 40.7 |
| **Operating Points C (◇)** |  |
|  | **C1 (NOCOS)** |
| Probability of survival | 91.3% |
| Predicted to survive & survived | 618 |
| Predicted to expire & expired | 388 |
| Predicted to survive & expired | 13 |
| Predicted to expire & survived | 1040 |
| Positive predictive value, % | 97.9 |
| Negative predictive value, % | 27.2 |

NOCOS = Northwell COVID-19 Survival; SOFA = Sequential Organ Failure Assessment.

**Appendix Table 4.** Determination of PaO_2_/FiO_2_ Score Based on SpO_2_ and FiO_2_ Values.

| **(SpO<sub>2</sub>/FiO<sub>2</sub>)\*100** | **PaO<sub>2</sub>/FiO<sub>2</sub> Score** |
| --- | --- |
| Null | 0 |
| ≥ 302.3 | 0 |
| Between 221.2 and 302.2 | 1 |
| Between 142.3 and 221.1 | 2 |
| Between 67 and 142.2 | 3 |
| < 67 | 4 |

